# COVID-19 Patients’ Symptoms: Gastrointestinal Presentations, Comorbidities and Outcomes in a Canadian Hospital Setting

**DOI:** 10.1101/2021.10.28.21265610

**Authors:** Hassan Brim, Michal Moshkovich, Melanie Figueiredo, Emily Hartung, Antonio Pizuorno, Lee Hill, Jelena Popov, Eyitope Olaide Awoyemi, Waliul I Khan, Gholamreza Oskrochi, Hassan Ashktorab, Nikhil Pai

## Abstract

**Background:** The Coronavirus disease 2019 (COVID-19) pandemic has had significant global impact. While public health interventions and universal health insurance has been credited with minimizing transmission rates in Canada relative to neighboring countries, significant morbidity and mortality have occurred nationwide. We sought to determine factors associated with differences in gastrointestinal outcomes in COVID-19 patients at a Canadian hospital.

**Methods:** We collected data from 192 hospital records of COVID-19 patients across seven Hamilton Health Sciences hospitals, a network of academic health centres serving one of the largest metropolitan areas in Canada. Statistical and correlative analysis of symptoms, comorbidities, and mortality were performed.

**Results:** There were 192 patients. The mean age was 57.6 years (SD=21.0). For patients who died (n=27, 14%), mean age was 79.2 years old (SD=10.6) versus 54 years for survivors (SD=20.1). There was a higher mortality among patients with older age (*p=*0.000), long hospital stay (*p=*0.004), male patients (*p=*0.032), and patients in nursing homes (*p=*0.000). Patients with dyspnea (*p=*0.028) and hypertension (*p=*0.004) were more likely to have a poor outcome. Laboratory test values that were significant in determining outcomes were an elevated INR (*p=*0.007) and elevated creatinine (*p=*0.000). Cough and hypertension were the most common symptom and comorbidity, respectively. Diarrhea was the most prevalent (14.5%) gastrointestinal symptom. Impaired liver function was related to negative outcome (LR 5.6; *p*=0.018).

**Conclusions:** In a Canadian cohort, elevated liver enzymes, prolonged INR and elevated creatinine were associated with poor prognosis. Hypertension was also linked to a higher likelihood of negative outcome.

**SUMMARY BOX:** *What is already known about this subject?:*

- The prevalence of gastrointestinal symptoms in COVID-19 patients across Canada is lacking
- Gastrointestinal manifestations of COVID-19 are well described, and longterm sequelae of gastrointestinal tract involvement are an ongoing concern

*What are the new findings?:*

- There was a significant prevalence of gastrointestinal symptoms in patients with a confirmed diagnosis of COVID-19 at one of the largest metropolitan regions across Canada
- Liver enzyme abnormalities were common in patients at diagnosis
- This report, over an 8-month period, represents the largest cohort of COVID-19 patients reported in Canada

*How might these results impact on clinical practice in the foreseeable future?:*

- Baseline gastrointestinal symptoms and laboratory abnormalities correlate with patient outcome in Canadian COVID-19 patients
- These results enhance our knowledge of the prevalence of gastrointestinal symptoms and laboratory abnormalities in Canadian patients and offer important baseline data for longitudinal studies in these patients
- Our findings increase our knowledge of the epidemiology of COVID-19 in Canada and allow future comparison with international data

## INTRODUCTION

Coronavirus Disease 2019 (COVID-19) was first reported in Wuhan, China in December 2019. This catastrophic illness has achieved pandemic status since the WHO declared the virus present in most countries by March 11, 2020, and has affected more than 92 million people with 2 million attributed deaths as of January 14^th^, 2020.^1^ On January 23^rd^, 2020, a 56-year old man presented to a hospital emergency department in Toronto, Canada, with new-onset fever and non-productive cough following return from Wuhan, China one day prior.^2^ This sentinel case marked the arrival of COVID-19 in Canada. Coronavirus is detected in both mid-turbinate and throat swabs by pancoronavirus RNA dependent RNA polymerase chain reaction (RdRp PCR), and confirmed as severe acute respiratory syndrome coronavirus 2 (SARS-CoV-2) by sequencing.^3^ As infection and hospitalization rates continue to rise, the need to identify disease-specific features and effectors within the Canadian population has become increasingly important.

As of February 12, 2021, Canada has confirmed 823,000 positive cases and more than 21,000 associated deaths.^4^ These numbers contrast with those from neighboring countries, such as Mexico and the United States, who are among the countries with the highest number of active cases and deaths. During the first wave of the pandemic (approximately January to July, 2020), COVID-19 appeared to exhibit a moderate case-rate and mortality rate nationwide. Factors which mitigated this initial impact included superior public health infrastructure, strict national lockdown measures, and universal healthcare access.

SARS-CoV-2 infects and damages the respiratory tract by adhering to angiotensin converting enzyme 2 (ACE2) receptors. The most common symptoms of SARS-CoV-2 infection include fever, cough, and myalgias.^5^ Although most patients experience mild to moderate disease, severe or critical disease requiring hospital admission may also develop and result in death in 1.5-3% of infected individuals.^6^

A growing body of literature has reported diarrhea, nausea, vomiting and other gastrointestinal (gastrointestinal) manifestations as potential symptoms of SARS-CoV-2 infection. Diarrhea is associated with other gastrointestinal tract infections, and it is not clear whether this symptom is specific to SARS-CoV-2 infection or a systemic response to the presence of intercurrent viral infections in general.^7^ The rate of any gastrointestinal symptom occurring at presentation or during illness in patients with COVID-19 has been reported between 16-50% across several studies.^8^ Two recent meta-analyses found the pooled prevalence of all gastrointestinal symptoms to be 17.6% with diarrhea, nausea, and vomiting being present in 7.7% and 7.8% patients with COVID-19 infection, respectively.^9,10^ It is worth noting that ACE2 receptor as well as the transmembrane protease serine 2 (TMPRSS2) enzyme that is required for SRAS-CoV-2 protein cleavage and virus entry are both expressed in the gastrointestinal tract and are likely to play some role in the gastrointestinal presentations.^11^

Canadian studies describing population demographics and clinical status of patients with COVID-19 are lacking. A better characterization is crucial to improve allocation of critical care resources and to improve our understanding of disease impacts with regional contexts. The aim of our analysis was to determine the rate of systemic and gastrointestinal symptoms, comorbidities, and laboratory test values in patients with COVID-19 across a densely populated region of Ontario, Canada’s highest populated province, and how these relate to outcomes in our patient population.

## METHODS

### Study design

We conducted a retrospective case series of all patients with COVID-19 who were seen across Hamilton Health Sciences (HHS), a network of hospitals, urgent care, rehabilitation, and long-term care centers. Seven institutions were included and all were located in Hamilton, Ontario, Canada. This study was approved by the Hamilton Integrated Research Ethics Board (Project #: 12862-C) and Howard University Office of Regulatory Research Compliance (IRB-12-CMED-76).

### Patients

Patients were screened between March, 2020 to October, 2020. HHS serves a catchment area of 2.3 million residents across southwestern Ontario and includes institutions dedicated to pediatric care, general adult medicine and subspecialty care, adult cancer care, long-term care and rehabilitation, and urgent care. This study included a wide spectrum of patients who received services at any of the affiliated hospitals and were found to be positive for SARS-CoV-2 by nasopharyngeal swab PCR during the screening period.

### Data Collection

Data were obtained from patient charts and electronic medical records where available. Demographic data, details of presenting and inpatient histories and examinations, laboratory data and clinical management were obtained from primary records. Patient demographics (age, sex, nursing home, known exposure, hospital admission, duration of symptoms, length of stay), symptoms (fever, dyspnea, cough, fatigue, myalgia, loss of taste, loss of appetite, abdominal pain, nausea, vomiting, diarrhea, dysphagia, gastrointestinal bleeding, pancreatitis, cholecystitis), underlying comorbidities (hypertension, cardiac disease, diabetes, obesity, smoking, alcohol intake, history of luminal gastrointestinal disease, liver disease, alcohol abuse, inflammatory bowel syndrome, gastrointestinal cancer, pancreatitis, gastroesophageal reflux disease or peptic ulcer disease), and laboratory data measured at baseline: international normalized ratio (INR) (0.8 - 1.2); total bilirubin (<21); direct bilirubin (<5); creatinine (male 60-110, female 49-90); C-reactive protein (<10.0); lactate dehydrogenase (120-250); fibrinogen (1.6-4.2); alanine aminotransferase (ALT) (male <50, female <35); albumin (42-50); platelets (130-400), creatine phosphokinase (CPK) (male less than 60 years old 45-250, male greater than 60 years old 40-200, female all ages 30-150); WBC (4.8-10.8); aspartate transaminase (AST) (male 18-54, female 18-34); alkaline phosphatase (ALP) (38-126). Data was omitted where unavailable. Results of chest radiographs, ultrasounds, and computerized tomography scans were also collected, including supportive and pharmacologic therapies received.

### Inclusion Criteria

Patients were included with a confirmed diagnosis of COVID-19, as reported by the Hamilton Regional Laboratory Medicine Program.^12^ We made no distinction with regards to sex, clinical manifestations, comorbidities, treatment, or outcome.

### Exclusion Criteria

The following exclusion criteria were adopted to filter out incomplete or ambiguous data: patients diagnosed with COVID-19 not confirmed by RT-PCR, patient charts with incomplete symptoms or comorbidities report, patients less than 19 years old.

### Statistical Analysis

Relationships to outcomes for demographic, clinical course, comorbidities, and laboratory data were assessed across our population. Correlation coefficients were calculated together with multivariate binary logistic regression analysis to establish associations with death as an outcome. SPSS version 26 (SPSS Inc., Chicago, IL, USA) was used for these analyses.^13^

## RESULTS

### i. Males over 50 years old were most common in our COVID-19 population

Our cohort consisted of 192 patients of which 51.4% were male (n=97) and 48.6% female (n=95). Our patients ranged between 19 and 95 years of age with a mean age of 57.6 years (SD=21.0). More than half of our cohort (50.5%, n=94) knew that they were exposed to others infected with SARS-CoV-2, and 21.2% of these (20 out of 94) patients were admitted to hospital. 51.5% (99 out of 192 patients) were seen in the Emergency Department. The length of stay in our patients’ cohort ranged from 0 to 90 days with a mean average of 11.5 days (SD=18.4) (Table 1).

**Table 1:**
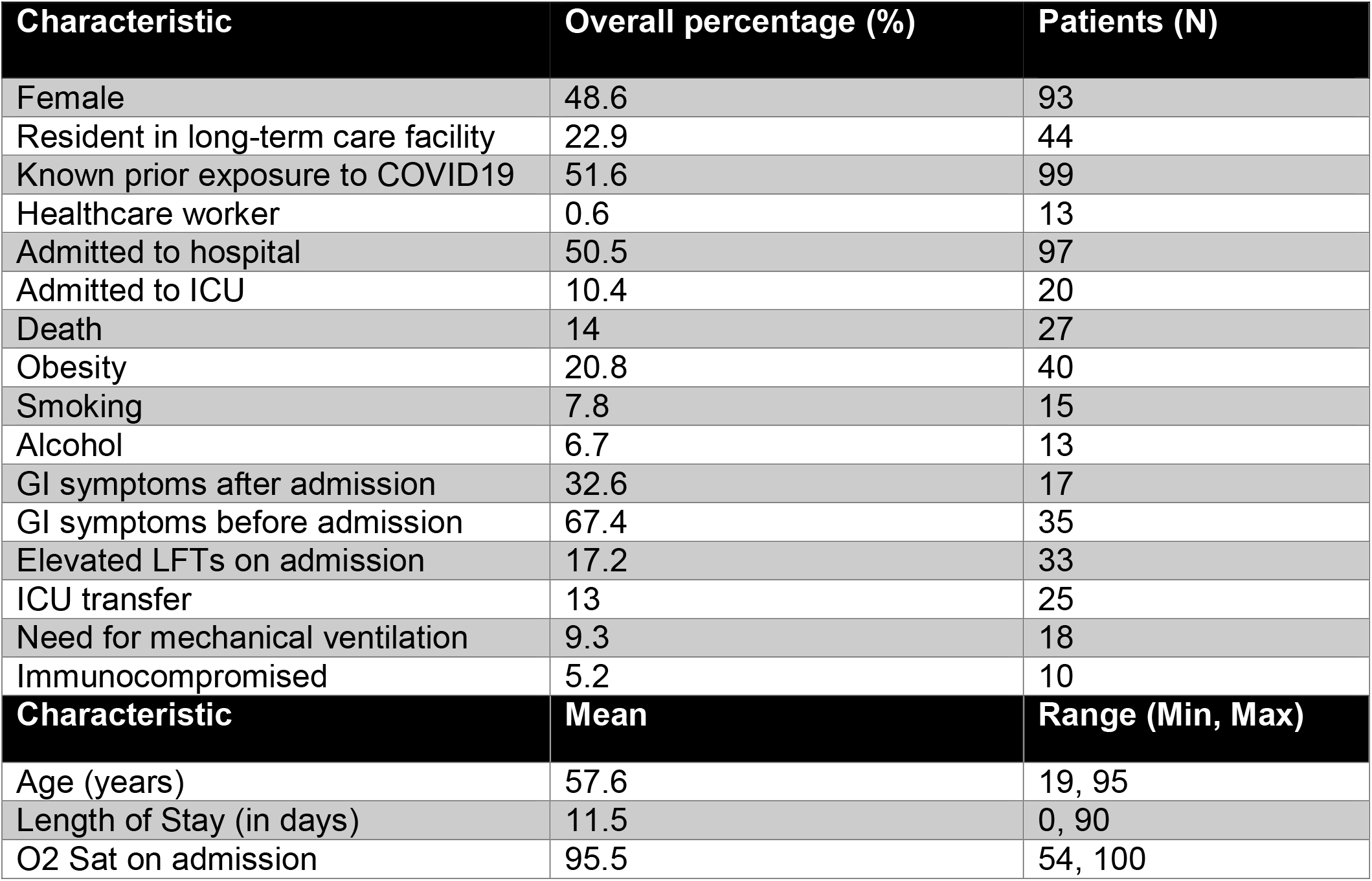
Patient demographics.

We recorded a total of 27 (14%) deaths during our review period. The mean age for those who survived was 54 years old (SD=20.1) and 79.2 years old (SD=10.6) for those who did not. Length of hospital stay also differed between those who survived (9.8 days; SD=17.9) versus for those who died (21.4 days; SD=19.0). Oxygen saturations were: 96.2% (SD=3.6) vs. 90.9% (SD=9.3) at admission, 94.5% (SD=2.2) VS. 94.5% (SD=2.7) at 24 hours, 94.8% (2.4) vs. 94% (2.5) at 48 hours, and 94.8% (2.8) vs. 93.5% (4.0) at 72 hours for those who survived versus for those who died, respectively (Table 1).

### ii. Symptoms and presentations

Overall, 55.2% (n=106) presented with cough, followed by dyspnea (47.1%, n=90), fatigue (36.4%, n=70), and fever (33.3%, n=64) (Table 2).

**Table 2:**
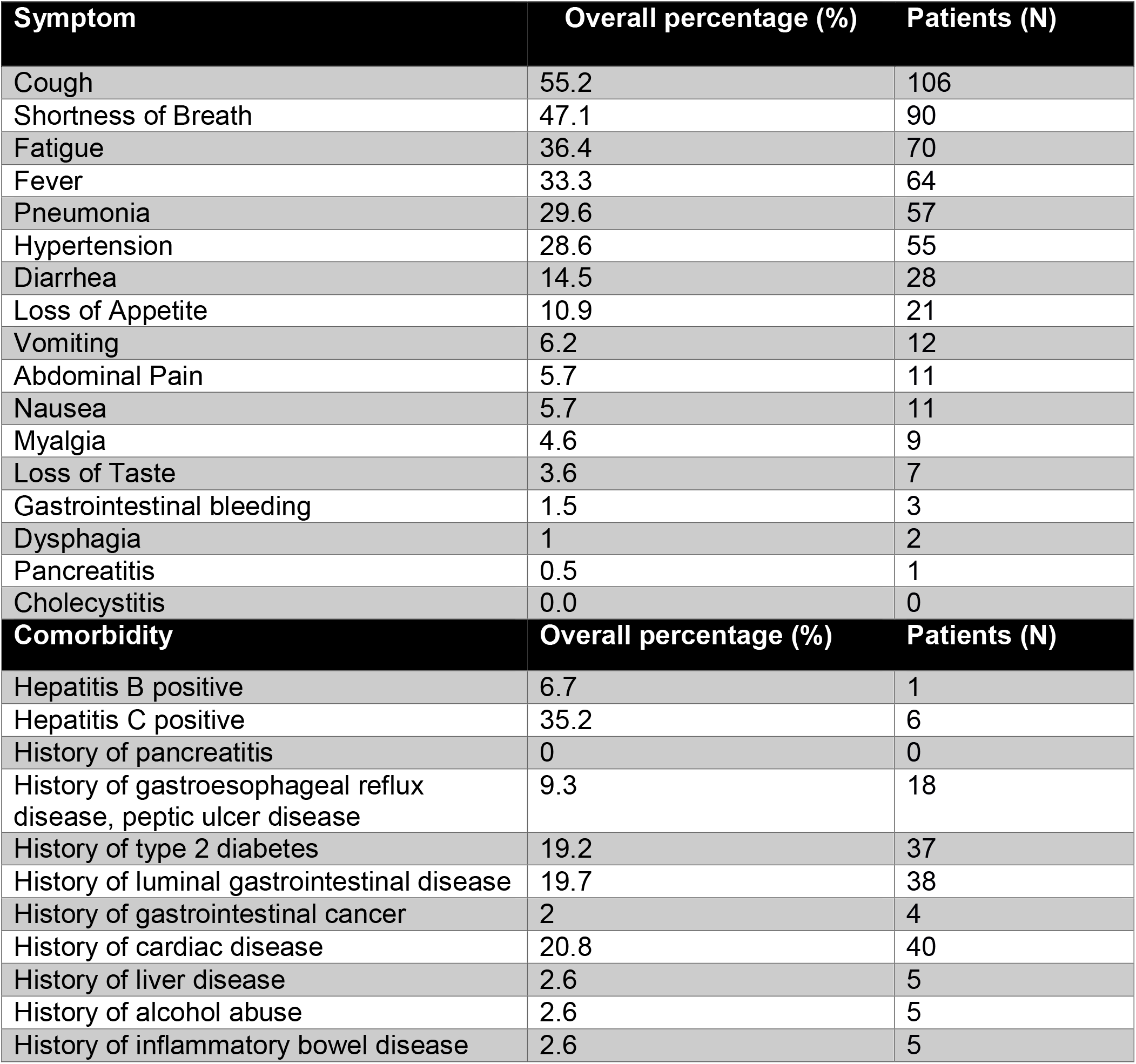
Presenting symptoms and associated comorbidities.

### iii. Multiple gastrointestinal symptoms

Gastrointestinal symptoms were prominent in our cohort (Table 2). The most common symptom was diarrhea (14.5%, n=28). Nausea and vomiting were reported in 5.7% (n=11) and 6.2% (n=12) cases, respectively. Abdominal pain was seen in 5.7% (n=11). Other reported gastrointestinal symptoms included: loss of appetite (10.9%, n=21), loss of taste (3.6%, n=7), and dysphagia (1.0%, n=2). Gastrointestinal bleeding, pancreatitis and cholecystitis were present in less than 1.5% of the cohort. In the 52 patients with gastrointestinal symptoms, 67.4% (n=35) had symptoms reported before admission, and the rest 32.6% (n=17) reported symptoms after admission.

### iv. Hypertension and diabetes mellitus were the most common chronic conditions

With respect to comorbidities, hypertension was reported in 28.6% (n=55), type 2 diabetes in 19.2% (n=37), and cardiovascular disease in 20.8% (n=40). Our patients were also carefully evaluated for concomitant gastrointestinal conditions. 19.7% (n=38) had a prior history of luminal gastrointestinal disease (including inflammatory bowel disease, diverticulitis), and 9.3% (n=18) had a history of gastroesophageal reflux disease or peptic ulcer disease. The least common presentations included a prior history of liver disease and inflammatory bowel disease, present in 2.6% each, and a history of gastrointestinal cancers, present in 2% (n=4) (Table 2).

### v. Laboratory tests revealed high levels of inflammatory markers

In our cohort, 80.3% (41 out of 51 patients) had elevated CRP levels, 62% (18 out of 29 patients) had elevated Lactate Dehydrogenase (LDH), and 18.1% (6 out of 13 patients) had elevated fibrinogen levels. Creatinine was high in 30.6% of patients (41 out of 134 patients), followed by elevated creatine phosphokinase (CPK) levels in 28.5% (12 out of 42 patients). Elevated creatinine was associated with 43.9% of patient deaths (18 out of 41 patients), among patients with high LDH levels, 33.3% died (n=6) and patients with low white blood cell (WBC) counts had a mortality rate of 35.5% (n=11) (Table 3).

**Table 3:**
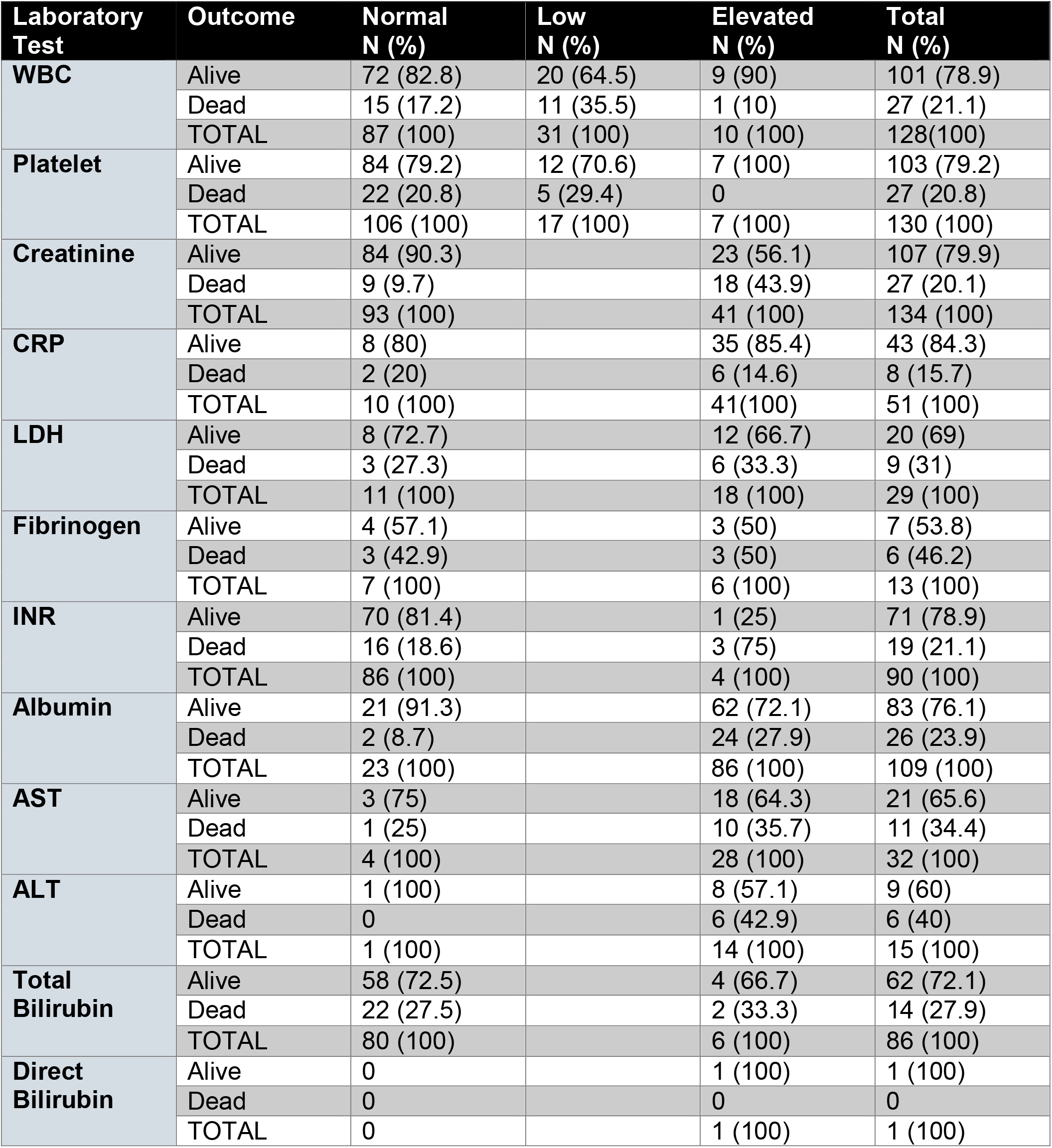
Laboratory tests and results by outcome.

### vi. Elevated ALT was common in the tested liver panel

The most frequent altered liver enzyme was alanine transaminase (ALT) with 93.3% (14 out of 15) with high levels. High levels of albumin occurred in 78.9% (86 out of 109). Elevated aspartate aminotransferase (AST) was reported in 87.5% (28 out of 32) when measured. Radiographic features of hepatic steatosis was present in 18.2% (2 out of 11 patients), and liver nodularity in 30.0% (3 out of 10 patients). Among deceased patients, the prevalence of elevated ALT was 42.9%, elevated AST 35.7%, elevated total bilirubin 33.3%, and elevated albumin 27.9% (Table 3).

### vii. Demographics were an important determinant of outcome

Older age (*p<*0.001), length of stay (*p=*0.004), and male sex (LR=4.7, *p=*0.030) were associated with death. Also, patients from nursing homes (LR=19.9, *p<*0.001), known exposure to an individual with COVID-19 (LR=5.1, *p=*0.024), and those who were admitted to hospital (LR=7.9, *p=*0.005) were more likely to die from their illness (Table 1).

### viii. Dyspnea and hypertension were related to mortality

Patients with dyspnea (LR=4.9; *p=*0.028) and hypertension (LR=7.6; *p=*0.006) were significant comorbidities among patients who died. None of the other clinical manifestations studied such as cough, fever, myalgia, diarrhea, abdominal pain, nausea, vomiting, or other frequent comorbidities such as diabetes mellitus, smoking, and alcohol consumption were significantly related to death (Table 2).

### ix. Inflammatory markers were not related to outcome

Although inflammation is generally high in COVID-19 patients, there was no correlation with outcome. Total bilirubin, direct bilirubin, CRP, LDH, fibrinogen, platelets, WBC, AST, ALT, and albumin were not related to outcome. Variables that could predict a poor outcome in our cohort were elevated INR (*p=*0.007) and elevated creatinine (*p<*0.001). Elevated liver function test at admission was highly predictive of negative outcome (*p=*0.001). Patients with a radiological diagnosis of pneumonia (*p<*0.001) and with an abnormal chest X ray (*p=*0.049) at baseline were more likely to have a negative outcome (Table 3).

## DISCUSSION

In this study, we comprehensively assessed characteristics and predictors of outcome of COVID-19 patients across a major metropolitan academic hospital network, consisting of seven major hospitals and treatment facilities located in Hamilton, Ontario, Canada. Our population included a catchment of approximately 2.3 million people, comprising the ninth largest metropolitan area in Canada.^14^ We found that baseline elevations in liver enzymes, creatinine, and INR are strong predictors of poor prognosis in Canadian patients receiving hospital care for COVID-19. The overall mortality rate for our cohort of 192 patients was 14% (n=27). Although gastrointestinal symptoms were not directly linked to outcome, they appeared to be highly prevalent in COVID-19 patients, which is noteworthy as SARS-CoV-2 virus persists within the gastrointestinal tract following clearance from the respiratory tract.^7^

In our cohort, patients who died were more likely to be older than 79.6 years (Table 1). Griesdale et al. reported a younger age of patients admitted to the ICU in Vancouver, Canada (69 years old).^15^ Patients who survived had a median age of 55 years old. It is worth noting that the median age of the Ontario population is 40.4 years old in 2020.^16^ Carignan et al. reported that patients with mild disease had a median age of 57.1 years old, which is similar to our cohort.^17^ Our findings that death affected older patients is in line with global data and further confirms that age-related fitness and associated immune response likely play a role in the protection against the deleterious effects of COVID-19.

Males and females were equally represented in our cohort. However, males were found to have a statistically significant increased risk of mortality. Griesdale et al. also reported that males were more likely to be admitted to intensive care unit (ICU) (67.5%) while Carignan et al. have reported a slight female predominance in mortality rates at 52.2%.^15,17^ The higher mortality rate among males has also been hypothesized to be due to potential protective immunologic functions on the X chromosome, or female sex-hormones.^18^

Cough was the most common symptom occurring in 55.2% of patients, with dyspnea (47.1%) and fever (33.3%) representing the next most common symptoms (Table 2). With the exception of dyspnea, these symptoms were not related to severity or mortality. This contrasts with data from O’Brien et al. who reported that in COVID-19 cases across Canada, the most common symptoms, in order of frequency were cough, fever, and chills. Dyspnea was among the sixth most common symptom.^19^

Gastrointestinal symptoms were also present in our cohort’s clinical manifestations. Diarrhea was present in 14.5% of patients, followed by vomiting (6.2%), nausea (5.7%), and abdominal pain (5.7%). Loss of appetite and taste occurred in 10.9% and 3.6% of patients, respectively. Although rare, few patients had gastrointestinal bleeding (1.5%) and pancreatitis (0.5%). Carignan et al. reported diarrhea in 44.8% of those with a positive SARS-CoV-2 test, while O’Brien et al. reported a rate of diarrhea of 23.6% in males and 27.8% in females.^17,19^ The overall incidence of acute gastrointestinal illness in the Canadian population in 2014-2015 was reported to be 25.6% [95% CI 18.1-34.9%].^20^ The presence of gastrointestinal symptoms points to how the effects of COVID-19 are not limited to the respiratory tract, but indeed spread systemically with different constellations of symptoms. The implications of gastrointestinal symptoms are also noteworthy. Patients with diarrhea have the possibility of transmitting the virus via fecal-oral routes and indeed, several investigators (including from Ottawa, Ontario) have used wastewater testing to capture the changing epidemiology of COVID-19 infection across metropolitan centers.^21^ This highlights the need for further investigations to fully assess the systemic effects of SARS-CoV-2 virus across the gastroenterological, neurological, and hematological systems.

Diabetes mellitus, hypertension, and obesity are the most common chronic conditions in Canada according to the Canadian Chronic Disease Surveillance System (CCDSS).^22^ The overall prevalence of obesity in the country is 26.9%, hypertension 25.5%, and type 2 diabetes 6.8%. Interestingly, although prevalent in our cohort, only hypertension was a risk factor for death in our study. This is in contrast with what has been cited by the Provincial Respiratory Surveillance Report from the province of Manitoba, which found that hypertension was present in 68% of hospitalized and 25% of non-hospitalized cases, while diabetes was reported in 45% of hospitalized versus 15% of non-hospitalized cases.^23^

Laboratory test values are important markers for general assessment of underlying determinants of observed symptoms and prognosis. Only elevated creatinine, elevated INR, and elevated ALT at admission were significantly associated with severe outcome and death in our cohort of COVID-19 patients. These findings corroborate with a study by Cheng et al. which reported that elevated creatinine and other indices of renal disease independently affected the outcome of hospitalized COVID-19 patients.^24^ Interestingly, in the study from Griesdale et al. including 117 patients, the only liver enzyme elevated in patients with COVID19 was AST. This study did not establish any clear relationships between laboratory values and patient outcomes like we did in our cohort.^15^

This was a retrospective study that followed patients over a period of eight months (March to October, 2020), during which clinical management protocols for COVID19 were actively evolving. Patient characteristics, laboratory values, and key demographic data were being actively identified from emerging publications worldwide, and our data reflects an evolution of how clinical record-keeping changed as new information became available. Nevertheless, we aimed to capture the most comprehensive data available, and most of our included variables reflected standardized data that is traditionally measured in all patients with acute illness or hospitalization.

In conclusion, age, male gender, elevated INR, creatinine, and liver enzymes were primary risk factors for poor outcome in our Canadian COVID-19 cohort of 192 patients from a major Canadian metropolitan region surrounding Hamilton, Ontario. Gastrointestinal symptoms were a prominent feature at presentation in our population, and while they did not appear to correlate directly with outcome, their presence reflects the multisystem nature of SARS-CoV-2 infection. Larger, multi-center studies are needed to offer a comprehensive picture of the COVID-19 infection across Canada.

## Data Availability

ll data produced in the present study are available upon reasonable request to the authors.

## ACKNOWLEDGEMENTS

The authors wish to thank Laura VanKuren, from Hamilton Health Sciences Health Information Technology Services, for her rapid assistance providing patient records to our study team to support data extraction.

